# Delayed first contact in a universally insured population: time to care and COVID-19 outcomes in Mexican primary care

**DOI:** 10.64898/2026.11.10.26367735

**Authors:** Víctor Junior Godínez-Vázquez, Efrén René Nieves Ruiz, Luis Fernando Razo Mendoza

## Abstract

Time from symptom onset to first care may be a modifiable determinant of COVID-19 outcomes, but it is rarely quantified in primary care. This retrospective cohort study included all 5,248 confirmed SARS-CoV-2 cases (RT-PCR or certified rapid antigen test) notified to the national surveillance system (SINOLAVE) by a large family medicine unit of the Mexican Social Security Institute (IMSS) in León, Mexico, between April 2020 and April 2021. Pandemic waves were classified using the official IMSS definition. Wave profiles were compared with chi-square tests and Bonferroni correction, and predictors of death were assessed by multivariable logistic regression, with sensitivity analyses adjusting for severity signs at first contact. Of the cases, 53.8% were female, mean age was 41.1 years, and 317 deaths (6.0%) were registered. Median time to first care was 3 days (interquartile range: 2–5). Case fatality was 3.0% among patients presenting within 3 days and 9.1% among those presenting later. Later presentation (> 3 days) was associated with death (adjusted odds ratio = 1.98; 95% confidence interval: 1.47–2.65), but the association was attenuated after adjustment for severity signs at first contact (adjusted odds ratio = 1.20; 95% confidence interval: 0.85–1.69), which were more frequent among later presenters (21.8% vs. 9.3%). From the first to the second wave, fever (76.1% to 56.9%), diabetes (22.8% to 9.6%), and case fatality (10.5% to 4.8%) decreased, whereas anosmia and dysgeusia increased (all p < 0.001). Surveillance data cannot distinguish clinical progression during delay from late presentation of already severe patients. Delay persisted despite universal entitlement to care, indicating that insurance coverage alone did not ensure timely presentation; prospective cohorts measuring severity at onset are needed.

## INTRODUCTION

The World Health Organization (WHO) declared COVID-19 a Public Health Emergency of International Concern on 30 January 2020^1,2^; the United States declared a nationwide public health emergency on 31 January 2020^3^, whereas Mexico declared a national health emergency on 30 March 2020^4^ with a delay of nearly 2 months. SARS-CoV-2, the third severe coronavirus outbreak after SARS (2002) and MERS (2012)^5^, uses the angiotensin-converting enzyme 2 (ACE2) receptor for cellular entry, with tropism for respiratory epithelium^6,7,8^. The later Omicron variant spread rapidly worldwide^9^. While hospital-based data are extensive, primary care surveillance data from Latin America remain underrepresented^10^. The Mexican Social Security Institute (IMSS) covers approximately 80 million beneficiaries, the largest social security system in Latin America^11^; Family Medicine Unit No. 47 in León is among its largest primary care facilities^12^. First contact facilities determine how quickly patients enter care, yet the prognostic weight of that delay is rarely quantified in primary care cohorts. Because all patients of this unit hold institutional entitlement to care, the cohort also allows delay to be examined in the absence of insurance-related financial barriers to access.

The objective of this study was to characterize the profile of confirmed cases and to assess whether time from symptom onset to first care was associated with mortality across the first two pandemic waves.

## METHODS

### Design, setting, and population

This retrospective cohort study was conducted at Family Medicine Unit No. 47 (IMSS), León, Guanajuato, Mexico, a high-volume first contact facility. All confirmed SARS-CoV-2 cases notified by the unit to the National Epidemiological Surveillance System (SINOLAVE) between 20 April 2020 and 24 April 2021 were included. A confirmed case was defined as a positive result on RT-PCR or on a certified rapid antigen test. Owing to molecular laboratory capacity constraints at the regional level, RT-PCR was performed on a subset of cases referred to the reference laboratory and served as a positivity index quality control validation of antigen-based confirmation; only COFEPRIS-certified and US FDA-authorized rapid antigen assays were accepted, and patients presenting results from noncertified assays were retested. WHO antigen performance thresholds applied (sensitivity ≥ 80%, specificity ≥ 97%)^13^. In SINOLAVE, 3,704 cases were registered as confirmed by RT-PCR and 1,544 by rapid antigen test. Pneumonia followed standard radiographic criteria^14^.

### Wave classification

Cases were assigned to pandemic waves by date of symptom onset using the official IMSS definition: Wave 1 through 10 October 2020 and Wave 2 from 11 October 2020 to 27 March 2021^15,16^. Seventy-six cases could not be assigned (75 with onset after 27 March 2021 and one with missing onset date) and were excluded from wave comparisons only. Predominant circulating lineages were ancestral/pre-Delta per national genomic surveillance; individual sequencing was not performed, and no causal variant associations are claimed.

### Exposure, outcomes, and statistical analysis

The primary exposure was time from symptom onset to first care at the unit (days), dichotomized at > 3 days for the adjusted model. The primary outcome was death registered in SINOLAVE as the discharge outcome, supported by a death certificate; secondary outcomes were radiographic pneumonia and hospital referral. Proportions are reported with Wilson 95% confidence intervals (95%CI). Wave 1 and Wave 2 were compared with Pearson’s chi-square test with Yates continuity correction and Bonferroni correction for 11 comparisons (significance threshold p < 0.0045), and mean age with Student’s t test.

Predictors of death were assessed by multivariable logistic regression including time to care, age, sex, diabetes, hypertension, and obesity, reporting adjusted odds ratios (OR) with 95%CI. The model included 5,245 cases; three were excluded (one negative interval, one interval > 30 days, and one missing date).

To assess whether the association could be explained by clinical severity at presentation, two sensitivity analyses were performed. First, the model was further adjusted for severity signs recorded at first contact: dyspnea, cyanosis, chest pain, clinical pneumonia, and radiographic pneumonia. Polypnea was recorded identically to cyanosis in all records and was not entered separately. Second, the primary model was refitted among cases without any of these signs at first contact. Vital signs, oxygen saturation, and laboratory results are not captured by SINOLAVE and could not be assessed. Analyses used Python (statsmodels and SciPy).

Generative AI (Claude, Anthropic) was used to assist with data reconciliation, drafting of Python scripts for statistical analysis and plotting (statsmodels, SciPy, matplotlib), and language editing. The conceptual diagram of competing explanations was produced in BioRender (biorender.com) from a layout specified by the authors using its AI-assisted generation tool. All scripts were reviewed and executed by the first author; tables and data figures were generated by these scripts directly from the surveillance data, and every statistic, table, and figure was verified against the source records. The authors take full responsibility for the content of this manuscript.

This study used anonymized secondary surveillance data under a consent waiver justified by the exclusive use of nonidentifiable records, and was approved by the Local Health Research Committee No. 1008 of IMSS (registration R-2021-1008-050; COFEPRIS 17 CI 11 020 063; CONBIOÉTICA 11 CEI 002 2018022).

## RESULTS

### Profile of confirmed cases

Among 5,248 confirmed cases, 2,822 (53.8%) were female. Mean age was 41.1 years (standard deviation, SD = 15.6; median 39; interquartile range, IQR: 29–51); 4,698 (89.5%) were aged 18–64 years and 473 (9.0%) were aged ≥ 65 years (Figure 1). The leading comorbidities were hypertension (19.6%; 95%CI: 18.5–20.7), obesity (16.8%; 95%CI: 15.8–17.8), and diabetes mellitus (12.5%; 95%CI: 11.6–13.4). The most frequent symptoms were headache (87.6%), cough (82.1%), and myalgia (69.6%); fever occurred in 61.0%. Most cases improved; 317 deaths were registered (case fatality 6.0%), radiographic pneumonia occurred in 1.3%, and 13.9% were referred to hospital care. Diabetes and COVID-19 form a bidirectional relationship, with admission hyperglycemia a recognized predictor of severity^17^.

**Figure 1.**
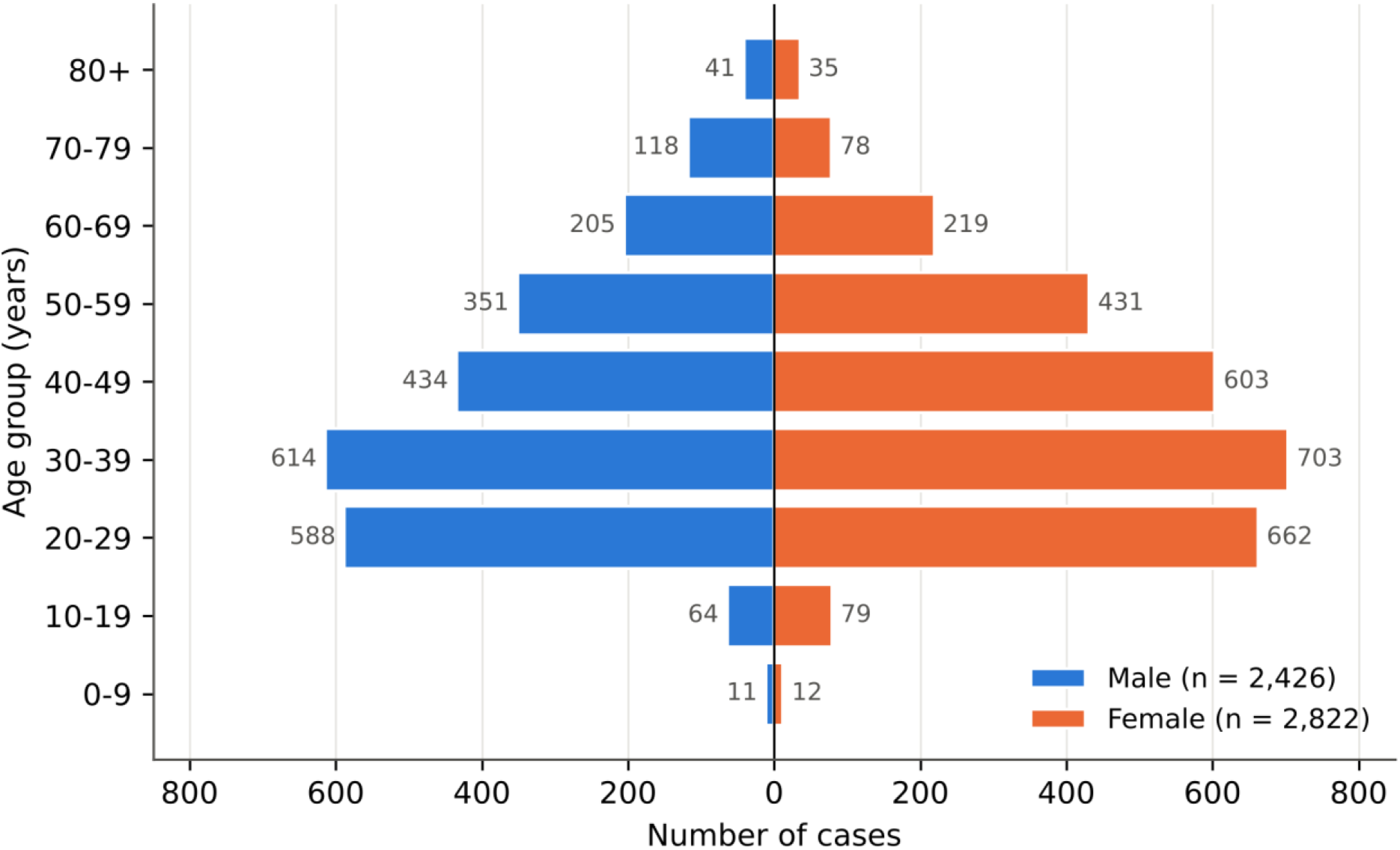
Age-sex distribution of confirmed COVID-19 cases (n = 5,248). Family Medicine Unit No. 47, Mexican Social Security Institute, León, Guanajuato, Mexico, April 2020–April 2021.

### Time to care and mortality

Median time from symptom onset to first care was 3 days (IQR: 2–5). Case fatality increased with delay: 3.0% (81/2,659) for presentation within 3 days versus 9.1% (236/2,586) thereafter (Figure 2). In the primary multivariable model, later presentation was associated with death (OR = 1.98; 95%CI: 1.47–2.65), together with age (OR = 1.11 per year), male sex (OR = 2.00), diabetes (OR = 2.08), and obesity (OR = 2.31); hypertension was not associated (OR = 0.92; 95%CI: 0.68–1.25) (Table 1).

**Table 1.** Multivariable logistic regression: factors associated with death (n = 5,245; 317 deaths)

| **Variable** | **Primary model OR (95%CI)** | **p** | **Severity-adjusted model OR (95%CI)** | **p** |
|---|---|---|---|---|
| Time to first care > 3 days (ref: 0–3 days) | 1.98 (1.47–2.65) | < 0.001 | 1.20 (0.85–1.69) | 0.299 |
| Age (per year) | 1.11 (1.10–1.12) | < 0.001 | 1.08 (1.06–1.09) | < 0.001 |
| Male sex (ref: female) | 2.00 (1.52–2.63) | < 0.001 | 1.87 (1.36–2.57) | < 0.001 |
| Diabetes mellitus | 2.08 (1.55–2.78) | < 0.001 | 1.70 (1.22–2.38) | 0.002 |
| Obesity | 2.31 (1.69–3.17) | < 0.001 | 1.87 (1.31–2.66) | 0.001 |
| Hypertension | 0.92 (0.68–1.25) | 0.606 | 1.03 (0.74–1.45) | 0.860 |
| **Severity signs at first contact** |  |  |  |  |
| Dyspnea | – |  | 15.23 (10.34–22.42) | < 0.001 |
| Cyanosis | – |  | 1.75 (1.00–3.04) | 0.049 |
| Chest pain | – |  | 1.00 (0.71–1.42) | 0.992 |
| Clinical pneumonia | – |  | 2.65 (1.62–4.32) | < 0.001 |
| Radiographic pneumonia | – |  | 0.68 (0.33–1.39) | 0.292 |

95%CI: 95% confidence interval; OR: odds ratio.
Polypnea was recorded identically to cyanosis in all records and was not entered separately. Among the 4,433 cases without severity signs at first contact (39 deaths), the OR for time to first care > 3 days was 1.53 (95%CI: 0.76–3.09).

**Figure 2.**
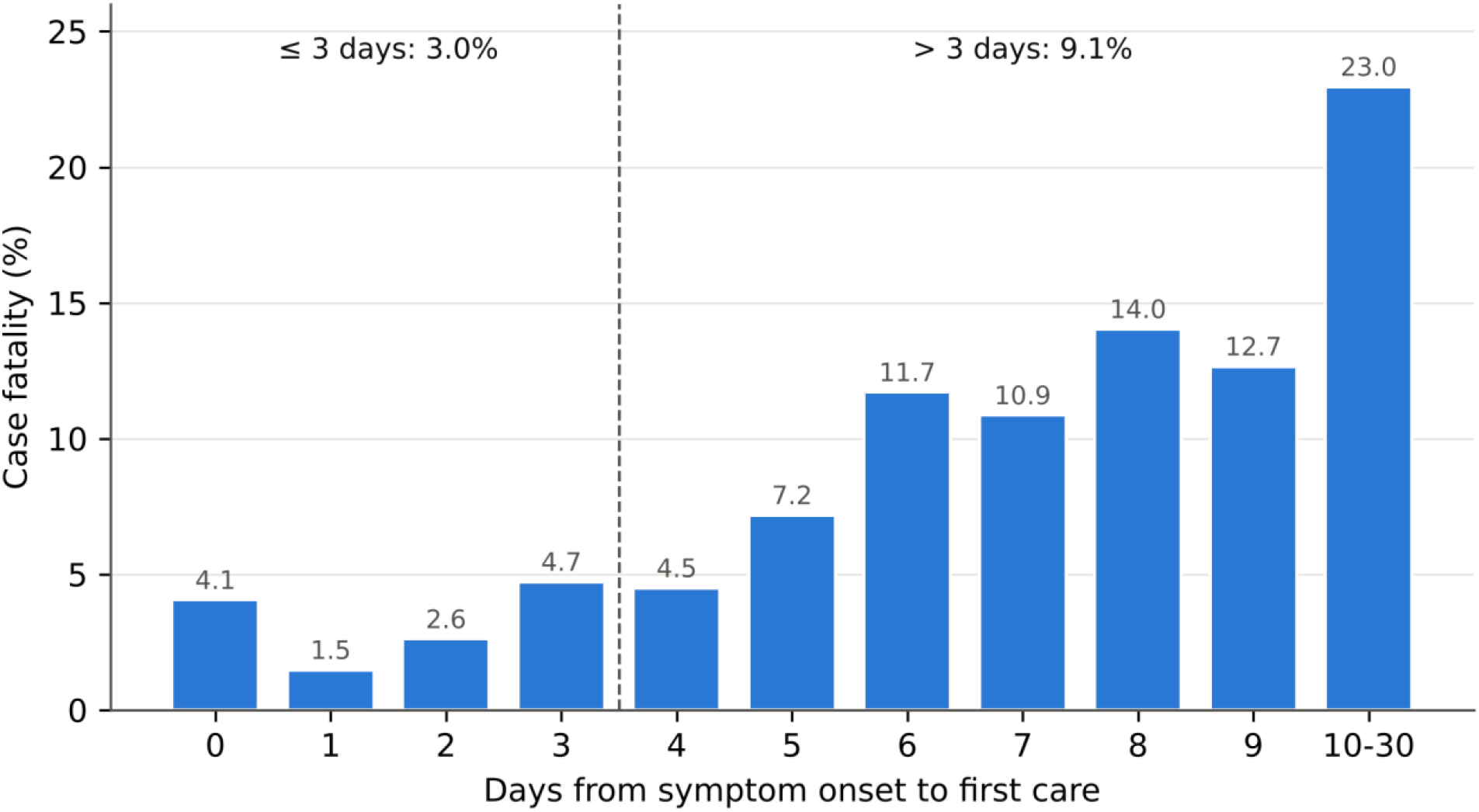
Case fatality by days from symptom onset to first care (n = 5,245; 317 deaths). León, Guanajuato, Mexico, April 2020–April 2021.

### Severity at first contact

At least one severity sign was recorded at first contact in 21.8% (565/2,586) of later presenters versus 9.3% (247/2,659) of those presenting within 3 days. After further adjustment for severity signs, the association between later presentation and death was attenuated and no longer statistically significant (OR = 1.20; 95%CI: 0.85–1.69); dyspnea (OR = 15.23; 95%CI: 10.34–22.42) and clinical pneumonia (OR = 2.65; 95%CI: 1.62–4.32) were the strongest predictors, while age, male sex, diabetes, and obesity remained associated (Table 1). Among the 4,433 cases without severity signs at first contact (39 deaths), the point estimate for later presentation was 1.53 (95%CI: 0.76–3.09).

### Evolution across the first two waves

Between Wave 1 (n = 1,167) and Wave 2 (n = 4,005), mean age decreased from 46.2 to 39.6 years (p < 0.001). Fever (76.1% to 56.9%), myalgia, chills, dyspnea, diabetes (22.8% to 9.6%), hypertension (32.0% to 16.1%), obesity, and case fatality (10.5% to 4.8%) decreased, whereas rhinorrhea (37.4% to 54.1%), anosmia (21.0% to 35.9%), and dysgeusia (21.3% to 34.9%) increased. All 11 differences remained significant after Bonferroni correction (p < 0.001) (Table 2).

**Table 2.** Clinical profile, comorbidities, and case fatality by pandemic wave.

| Variable | Wave 1 (n = 1,167) n (%) | Wave 2 (n = 4,005) n (%) | Chi-square | p-value |
|---|---|---|---|---|
| **Symptoms** |  |  |  |  |
| Fever | 888 (76.1) | 2,280 (56.9) | 139.0 | < 0.001 |
| Myalgia | 903 (77.4) | 2,708 (67.6) | 40.4 | < 0.001 |
| Chills | 689 (59.0) | 1,945 (48.6) | 39.3 | < 0.001 |
| Dyspnea | 228 (19.5) | 458 (11.4) | 50.9 | < 0.001 |
| Rhinorrhea | 437 (37.4) | 2,168 (54.1) | 100.0 | < 0.001 |
| Anosmia | 245 (21.0) | 1,438 (35.9) | 90.9 | < 0.001 |
| Dysgeusia | 249 (21.3) | 1,396 (34.9) | 75.5 | < 0.001 |
| **Comorbidities** |  |  |  |  |
| Diabetes mellitus | 266 (22.8) | 385 (9.6) | 141.5 | < 0.001 |
| Hypertension | 373 (32.0) | 644 (16.1) | 143.3 | < 0.001 |
| Obesity | 290 (24.9) | 581 (14.5) | 68.3 | < 0.001 |
| **Outcome** |  |  |  |  |
| Death | 123 (10.5) | 192 (4.8) | 51.2 | < 0.001 |

Pearson chi-square with Yates continuity correction, 1 degree of freedom; Bonferroni threshold 0.05/11 = 0.0045. Mean age: Wave 1 46.2 (SD 16.1) vs. Wave 2 39.6 (SD 15.1) years, p < 0.001.
Wave 1: symptom onset through 10 October 2020; Wave 2: 11 October 2020 to 27 March 2021 (official IMSS definition). 76 cases with onset outside both waves or missing onset were excluded from this comparison.

## DISCUSSION

In 5,248 confirmed cases at a large primary care unit, later presentation to care was associated with mortality, case fatality tripled beyond three days from onset, and the clinical profile shifted significantly between the first two waves. The association was largely explained by greater severity at first contact, which was more than twice as frequent among later presenters.

These data cannot distinguish two explanations (Figure 3). Clinical worsening may lie on the causal pathway between delay and death, in which case adjustment for severity at first contact removes part of the effect of delay itself; alternatively, patients already severe at onset may have presented later. Under the first explanation, time to care remains potentially modifiable through community messaging, triage access, and lowered thresholds for first contact; under the second, earlier care would not by itself change outcomes. Resolving this requires prospective cohorts that measure severity at symptom onset and at first contact, with formal mediation analysis.

**Figure 3.**
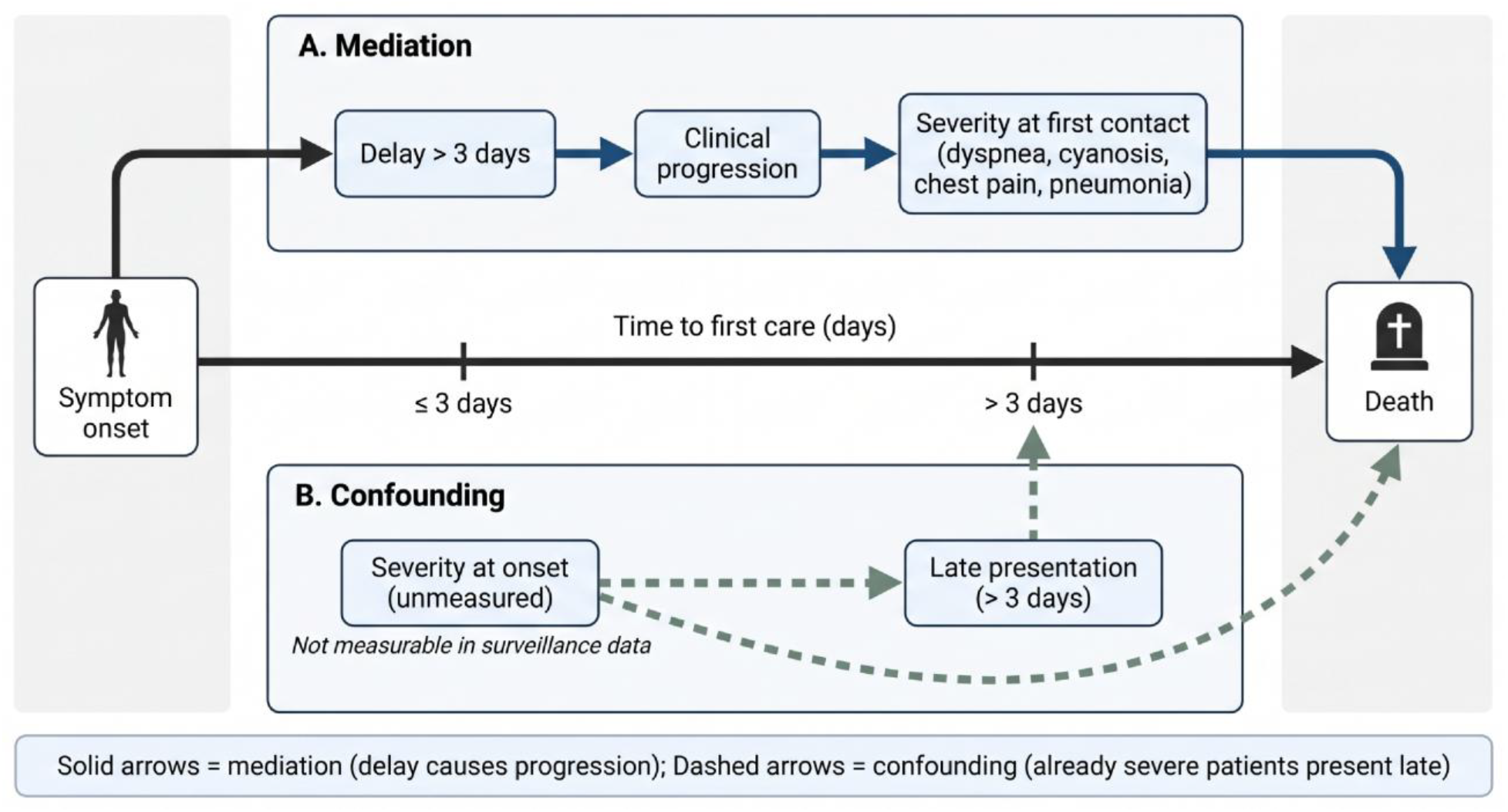
Competing explanations for the association between time to first care and COVID-19 mortality: clinical progression during delay (A, mediation) versus later presentation of patients already severe at onset (B, confounding).

The female predominance (53.8%) contrasts with hospital cohorts showing male predominance^18^ and aligns with community and long-term care data^19^, possibly reflecting differential care seeking. The cardiometabolic burden was lower than in hospitalized cohorts such as New York City^20^, as expected in a first contact population, yet reflects Mexico’s chronic disease epidemic. The shift from a febrile, comorbid, older Wave 1 profile toward a younger profile with more anosmia and dysgeusia in Wave 2 is consistent with evolving circulating lineages and exposure documented nationally^15^. The study window is essentially prevaccination^21^, providing a baseline profile; the cohort’s neurological and systemic symptom burden aligns with manifestations later reported in long COVID^22,23,24,25^, for which primary care is well positioned for longitudinal surveillance.

Two strategies, suppression and mitigation, have framed pandemic management^26^. Delayed presentation in an insured Mexican population suggests that access delay operates even absent the insurance and language barriers documented among US Hispanics^27^, pointing to determinants beyond coverage alone; culturally patterned care seeking behaviours (familismo, personalismo, jerarquismo, espiritismo) were not measured here and warrant dedicated mixed methods study. Prospective studies of the mediation question are particularly relevant for Hispanic populations in Mexico and the United States, where such care seeking patterns may prolong time to care. This patient-level finding stands in asymmetric relation to system-level delay documented during Mexico’s national response, which a case analysis attributed in part to underestimation of risk and limited coordination across government^28^. First contact facilities absorbed the clinical consequences of delayed presentation.

A specific mechanism warranting investigation is the role of health misinformation in delaying care seeking; the COVID-19 infodemic has been modelled epidemiologically, with surveillance ideally triggered before misinformation propagates widely^29^. Whether such dynamics contributed to the delays observed here is untested and merits study integrating clinical, perceptual, and information exposure data.

If time to care is a modifiable contributor to mortality, then delayed presentation is not solely a clinical variable but a marker of differential access to care. Framing avoidable delay through a health equity lens implies that preparedness should address the social determinants shaping who reaches care in time, rather than only the biological susceptibility that clinical management targets. In this cohort, insurance-related financial barriers were absent by design: all patients held institutional entitlement to care. Delay nonetheless persisted. Its determinants were not measured here and cannot be inferred from these data: neither predisposing factors and perceived need, nor enabling resources other than financing, such as travel distance, appointment availability, unit saturation, and the indirect costs of absence from work, could be assessed. Identifying which of these predominate is the priority for future research in pandemic preparedness.

This study has limitations. Deaths were those registered by the unit in SINOLAVE; outcomes of referred cases were only partially ascertained, so case fatality represents registered first contact mortality rather than total mortality, and deaths after referral may be undercounted. Severity was available only as signs recorded at first contact; vital signs, oxygen saturation, and laboratory results were not captured, and severity at symptom onset was not measured. Patients who died before reaching first contact were not captured, which may bias the delay-mortality association; time to care relied on patient-reported onset; variant attribution is inferential; unmeasured confounders, including socioeconomic status, may influence the delay-mortality association; and this single-unit cohort requires multisite validation.

## CONCLUSIONS

At a large primary care unit in Mexico, delayed presentation was associated with registered COVID-19 mortality, but the association was largely explained by greater severity at first contact, and the clinical profile shifted significantly between the first two pandemic waves. Whether delay causes clinical progression or reflects late presentation of patients already severe cannot be determined from surveillance data. Delay persisted in a population with universal entitlement to care, indicating that insurance coverage alone did not ensure timely presentation. Prospective cohorts measuring severity at symptom onset and at first contact are needed to establish whether reducing time to first care is an effective preparedness target for primary care.

## Data Availability

The data underlying this study are anonymized surveillance records from the National Epidemiological Surveillance System (SINOLAVE) and are held by the Instituto Mexicano del Seguro Social. They are not publicly available owing to institutional restrictions, but are available from the authors upon reasonable request and with authorization from the Instituto Mexicano del Seguro Social.

## ACKNOWLEDGMENTS

This study received no funding.

## Notes

### Competing Interest Statement

The authors have declared no competing interest.

### Author Declarations

The Health Research Committee No. 1008 (Comite Local de Investigacion en Salud 1008, Unidad de Medicina Familiar No. 53) of the Instituto Mexicano del Seguro Social (Mexican Social Security Institute) gave ethical approval for this work (institutional registration R-2021-1008-050; COFEPRIS 17 CI 11 020 063; CONBIOETICA 11 CEI 002 2018022).

